# Characterization of the genetic relationship between the domains of sleep and circadian-related behaviors with substance use phenotypes

**DOI:** 10.1101/2021.12.09.21267547

**Authors:** Alexander S. Hatoum, Evan A. Winiger, Claire L. Morrison, Emma C. Johnson, Arpana Agrawal

## Abstract

Sleep problems and substance use frequently cooccur. While substance use can often manifest as specific sleep deficits, genetic pleiotropy could also explain part of the relationship between sleep and substance use. Here we assess the genetic overlap between substance use behaviors and both sleep and circadian-related activity measures by deriving genetic clusters between these domains and testing processes of causality vs. horizontal pleiotropy using the largest publicly available genome-wide summary statistics of substance use behaviors (N= 79,729 - 632,802) and sleep/activity phenotypes/endophenotypes to date (N=85,502 - 449,734). We found 31 genetic correlations between substance use and sleep/activity measures after Bonferroni correction. Two specific genetic clusters explained our patterns of overlap. Genes associated with tobacco use severity (age of first regular tobacco use and smoking cessation) share overlap with elements of sleep health (sleep duration, sleep efficiency, and chronotype). Substance consumption (drinks per day and cigarettes per day) and problematic substance use behaviors (cannabis use disorder, opioid use disorder, and problematic alcohol use) clustered strongly with problematic measures of sleep (insomnia, self-reported short sleep duration, increased number of sleep episodes, increased sleep duration variability, diurnal inactivity) as well as measures of circadian-related activity (L5, M10, and sleep midpoint). Latent causal variable analyses determined that horizontal pleiotropy (rather than causality) underlies a majority of the associations between substance use and sleep/circadian related measures, except one plausible genetically causal relationship for opioid use disorder on self-reported long sleep duration. Results indeed show significant genetic overlap between substance use and sleep/circadian-related activity measures.

## Introduction

Sleep disturbances are one of the most common complaints in substance use treatment, ^1, 2^ and there is substantial comorbidity between substance use disorders (SUD) and sleep disorders ^3, 4^. Attempting to quit using substances and the consequent cravings and withdrawal are often associated with sleep disturbances and these sleep deficits can in turn be reciprocally linked to substance use relapse ^5^. For instance, sleep difficulties and insomnia are common clinical features of withdrawal from alcohol, nicotine, and cannabis ^6^. Further complicating this relationship, common substances, such as alcohol and cannabis, are often used to self-medicate sleep issues, with evidence that cannabis ^7–9^ and alcohol ^8–11^ are frequently considered sleep aids despite strong cross-sectional evidence demonstrating positive correlations between increased use of many common substances and sleep issues ^5^. Relatedly, tobacco is often used as a stress and tension reliever ^12, 13^ but tobacco use before sleep is also associated with sleep disturbances^14^.

Research suggests that circadian mechanisms contribute to the association between sleep issues and substance use/abuse, and that substance use may also impact circadian rhythms, thus further disturbing sleep ^15^. While circadian rhythm is a multi-faceted biological construct, it is often operationally defined as chronotype, a preference between either evening or morning activity, wake-up, and bedtime ^16^. An evening chronotype has been associated with higher alcohol ^17–19^, tobacco ^18, 20, 21^, and cannabis use ^19, 20, 22^. In addition to the standard self-report measure of chronotype, activity-related proxies (measured via actigraphy or accelerometer) can function as objective circadian-related measures ^23–26^. Research on the relationship between circadian-related activity measures and substance use is scarce ^27^ and is yet to be fully understood.

Sleep and substance use behaviors seem to interact dynamically. There is evidence of a bi-directional relationship between the domains of substance use and sleep deficits particularly during development there is indication that early sleep problems and an evening chronotype predict later substance use ^19, 28–35^ and both early substance use and problematic substance use predicts later sleep issues ^28, 29, 36–42^. Reconciling these cross-sectional associations and bidirectional patterns, it is likely that a third variable, such as genetic pleiotropy, is driving trait-like manifestations of sleep and substance use comorbidity.

Here, we focus on trait like (rather than state-like) overlap in sleep and substance use by utilizing statistical genetics methods. Prior genetic studies implicate shared genetic influences on sleep and substance use and misuse as a likely contributor to their comorbidity. Twin studies focused on sleep/circadian-related outcomes and substance use are scarce but a few have found genetic correlations between regular cannabis use and both short sleep duration and insomnia^41, 42^ as well as genetic correlations between an evening chronotype with alcohol quantity and increased binge drinking ^43^. Modern genome-wide association studies (GWASs) studies have found genetic correlations amongst subjective sleep deficits and common substance use behaviors including tobacco behaviors such as smoking initiation, smoking cessation, and cigarettes per day ^44, 45^, alcohol behaviors via scores on The Alcohol Use Disorders Identification Test (AUDIT) ^46^, Opioid Use Disorder ^47^, and cannabis behaviors such as lifetime cannabis use and Cannabis Use Disorder ^48^. GWASs focused on chronotype/circadian-related activities and substance use are sparse. Genetic correlations have been found between an evening chronotype and lifetime cannabis use ^48^, but the relationship between the multitude of available substance use and both chronotype and circadian-related activity measures has yet to be explored. Still, the current limited genetically informed results imply that the genes that could be contributing to sleep deficits and circadian-related factors such as chronotype might contribute to substance use and misuse as well or vice versa.

Genetic studies can also be used to inform causality. A prior study explored the pairwise associations between sleep and substance use disorders using Mendelian randomization, finding that insomnia had a potential positive causal influence on smoking, alcohol dependence, and cannabis initiation, while smoking initiation may be causal for insomnia ^49^. However, the effects of insomnia on both alcohol dependence and cannabis initiation were partially driven by pleiotropic SNPs (rather than direct causality), suggesting caution in inferring these results as causal and the possibility of a shared genetic liability. Thus, a perspective incorporating pleiotropy is needed.

Finally, there has yet to be a comprehensive genomic study to include multitude of alcohol, tobacco, cannabis, and opioid use behaviors as well as both a collection of subjective and objective/endophenotype sleep/circadian-related measures. Sleep itself is a highly heterogenous set of behaviors, encompassing many subdomains that relate differentially. Substance use and use disorder may interfere with sleep generally, or with sleep efficiency, duration, circadian rhythm, or the sleep cycle to produce sleep problems. Further, different substances may show unique relationships with any one domain of sleep specifically. Therefore, it is important to explore subdomains of substance use, misuse, and sleep behaviors in order to localize the causes of such deficits.

No study has comprehensively looked across the relationships of substance use behaviors and sleep/activity measures with the goal of defining genetic clusters of sleep/circadian-related behaviors and substance use traits. Therefore, the goals of the current study were to:

1) Investigate the shared genetics between a multitude of substance use and sleep/activity measures. 2) Determine if genetic causality or genetic pleiotropy are responsible for the genetic relationships discovered. 3) Analyze genetic clusters amongst the domains of substance use and sleep/activity.

## Methods

### Measures

#### Subjective and Objective Sleep and Circadian-related Measures

Our analyses used summary statistics from several large scale GWAS focused on self-report sleep phenotypes and objective accelerometer derived sleep/activity phenotypes/endophenotypes. Self-report sleep phenotypes included insomnia, chronotype, self-report sleep duration, self-report short sleep duration, and self-report long sleep duration ^50–53^. Objective accelerometer derived sleep/circadian-related activity phenotypes/endophenotypes included sleep duration, standard deviation of sleep duration (a measure of sleep variability), sleep efficiency, number of sleep episodes, diurnal inactivity (inactive states such as napping and wakeful rest), sleep midpoint (a proxy of chronotype ^54, 55^), least active 5 hours of the day (L5 timing, indication of a preference for going to bed earlier or later in the day ^23^), and most active 10 hours of the day (M10 timing, indication of if a person is most active earlier or later in the day^23^) ^56^. Diurnal inactivity can be conceptualized as a sleep problem due to increased naps/rest/inactivity that are a compensatory behavior for non restful sleep ^57^. L5, M10, and sleep midpoint will be conceptualized as activity measures that are circadian-related proxy measures. Table 1 details sleep/activity trait in terms of type of measure, measure construct, coding, and discovery sample.

**Table 1.** Trait, type of measure, measure construct, coding, and discovery sample for all substance use and sleep and activity phenotypes/endophenotypes.

| **Trait** | **Type of measure** | **Discovery Sample** | **Measure construct** | **Coding** |
| --- | --- | --- | --- | --- |
| Insomnia<br>(Jansen et al. 2019) | Self-report | UK Biobank | Insomnia complaints were assessed by asking: “Do you have trouble falling asleep at night or do you wake up in the middle of the night?” Insomnia cases were defined as participants who answered this question with “usually”, while participants answering “never/rarely” or “sometimes” were defined as controls. | $n = 386,533$ |
| Chronotype<br>(Jones et al. 2019) | Self-report | UK Biobank | Self-reported measure phrased as “Do you consider yourself to be?” with one of six possible answers: “Definitely a ‘morning’ person”, “More a ‘morning’ than ‘evening’ person”, “More an ‘evening’ than a ‘morning’ person”, “Definitely an ‘evening’ person”, “Do not know” or “Prefer not to answer”. | $n = 449,734$<br>Definitely morning, 2, $n = 107,555$<br>More morning than evening, 1, $n = 144,731$<br>Don’t know, 0, $n = 46,538$<br>More evening than morning, -1, $n = 115,090$<br>Definitely evening, -2, $n = 35,818$ |
| Self-report sleep duration<br>(Dashti et al. 2019) | Self-report | UK Biobank | A continuous variable categorized as short (6h or less), normal (7–8h), or long (9h or more) sleep duration. Responses of less than 3h or more than 18h excluded. | $n = 446,118$ |
| Self-report short sleep duration<br>(Dashti et al. 2019) | Self-report | UK Biobank | Short sleep duration (<7h) compared to 7–8h sleep duration. | $n = 446,118$; short (<7h; $n = 106,192$ cases) compared to 7–8h sleep duration ($n = 305,742$ controls) |
| Self-report long sleep duration<br>(Dashti et al. (2019) | Self-report | UK Biobank | Long sleep duration ($\geq 9$h) compared to 7–8h sleep duration. | $n = 446,118$; long ($\geq 9$h; $n = 34,184$ cases) compared to 7–8h sleep duration ($n = 305,742$ controls) |
| L5 timing<br>(Jones et al. 2019) | Accelerometer | UK Biobank | The midpoint of the least active 5h of each day. This measure implies whether someone prefers to go to bed earlier or later in the day. | $n = 85,830$ |
| M10 timing<br>(Jones et al. 2019) | Accelerometer | UK Biobank | The most active 10□h of each day. This measure implies whether a person is most active earlier or later in the day. | $n□=□85,723$ |
| Sleep duration in hours<br>(Jones et al. 2019) | Accelerometer | UK Biobank | Summed duration of all sleep episodes. | $n□=□85,068$ |
| Sleep duration in hours (standard deviation)<br>(Jones et al. 2019) | Accelerometer | UK Biobank | Standard deviation of the summed duration of all sleep episodes. | $n□=□85,068$ |
| Sleep midpoint<br>(Jones et al. 2019) | Accelerometer | UK Biobank | The midpoint between the start of the first detected sleep episode and the end of the last sleep episode. | $n□=□85,502$ |
| Sleep efficiency (%)<br>(Jones et al. 2019) | Accelerometer | UK Biobank | sleep duration divided by the time between the start and end of the first and last nocturnal inactivity period. | $n□=□85,502$ |
| Number of sleep episodes<br>(Jones et al. 2019) | Accelerometer | UK Biobank | Periods of at least 5□min with no change larger than 5° associated with the z-axis of the activity-monitor. | $n□=□85,502$ |
| Diurnal inactivity<br>(Jones et al. 2019) | Accelerometer | UK Biobank | The total daily duration of estimated bouts of inactivity that fell outside of the sleep period time window. This measure captures very inactive states such as napping and wakeful rest but not inactivity such as sitting and reading or watching television. | $n□=□85,502$ |
| Alcoholic drinks per week (DPW)<br>(Liu et al., 2019) | Self-report | Meta-analysis performed from, Add Health, Avon Longitudinal Study of Parents and Children, Atherosclerosis Risk in Communities, Biobank Japan Project, The Barrett's and Esophageal Adenocarcinoma. Genetic Susceptibility Study, Brisbane Longitudinal Twin Study, Center on Antisocial Drug Dependence, Collaborative Genetic Study of Nicotine Dependence, Genetics of Chronic Obstructive Pulmonary Disease, deCODE Genetics/AMGEN, Inc., Estonian Genome Center, Electronic Medical Records and Genomics, Finnish Twin | Average number of drinks a participant reported drinking each week. | $n = 537,349$ |
|  |  | Cohort, Framingham Heart Study, Genetic Epidemiology Research in Adult Health and Aging, Genes for Good, The Nord-Trøndelag Health Study, Health and Retirement Study, Minnesota Center for Twin and Family Research, Multi-Ethnic Study of Atherosclerosis, Metabolic Syndrome in Men, Netherlands Study on Cognition, Environment and Genes, Nurses' Health Study, Nurses' Health Study II, and Health Professionals' Follow-up Study, The National Institute of Neurological Disorders and Stroke Genetics Network, Netherlands Twin Register, Australian Twin-Family Studies on Nicotine and Alcohol Genetics, SardiNIA project), UK Biobank, and Women's Health Initiative |  |  |
| Problematic alcohol use (PAU)<br>(Zhou et al., 2020) | Self-report | Million Veteran Program, UK Biobank, Psychiatric Genomics Consortium Substance Use Disorders working group AUD and AUDIT-P, European-ancestry individuals | DSM-IV alcohol dependence (AD), AUD via the International Classification of Diseases (ICD) codes, Alcohol Use Disorders Identification Test–Problems (AUDIT-P). | $n = 435,563$ |
| Lifetime tobacco use<br>(Liu et al., 2019) | Self-report | Meta-analysis performed from 23&me, Add Health, Avon Longitudinal Study of Parents and Children, Atherosclerosis Risk in Communities, Biobank Japan Project, The Barrett's and Esophageal Adenocarcinoma Genetic Susceptibility Study, Brisbane Longitudinal Twin Study, Center on Antisocial Drug Dependence, Collaborative Genetic Study of Nicotine Dependence, Genetics of Chronic Obstructive Pulmonary Disease, deCODE Genetics/AMGEN, Inc., Estonian Genome Center, Electronic Medical Records and Genomics, Finnish Twin Cohort, Framingham Heart Study, Genetic Epidemiology Research in Adult Health and Aging, Genes for Good, The Nord-Trøndelag Health Study, Health and Retirement Study, Minnesota Center for Twin and Family Research, Multi-Ethnic | Reporting ever being a regular smoker in their life (current or former) coded as “2” and any participant who reported never being a regular smoker in their life coded as “1”. | $n = 632,802$ |
|  |  | Study of Atherosclerosis, Metabolic Syndrome in Men, Netherlands Study on Cognition, Environment and Genes, Nurses' Health Study, Nurses' Health Study II, and Health Professionals' Follow-up Study, The National Institute of Neurological Disorders and Stroke Genetics Network, Netherlands Twin Register, Australian Twin-Family Studies on Nicotine and Alcohol Genetics, SardiNIA project), UK Biobank, and Women's Health Initiative |  |  |
| Age of first becoming a regular smoker<br>(Liu et al., 2019) | Self-report | Meta-analysis performed from, Add Health, Avon Longitudinal Study of Parents and Children, Atherosclerosis Risk in Communities, Biobank Japan Project, The Barrett's and Esophageal Adenocarcinoma Genetic Susceptibility Study, Brisbane Longitudinal Twin Study, Center on Antisocial Drug Dependence, Collaborative Genetic Study of Nicotine Dependence, Genetics of Chronic Obstructive Pulmonary Disease, deCODE Genetics/AMGEN, Inc., Estonian Genome Center, Electronic Medical Records and Genomics, Finnish Twin Cohort, Framingham Heart Study, Genetic Epidemiology Research in Adult Health and Aging, Genes for Good, The Nord-Trøndelag Health Study, Health and Retirement Study, Minnesota Center for Twin and Family Research, Multi-Ethnic Study of Atherosclerosis, Metabolic Syndrome in Men, Netherlands Study on Cognition, Environment and Genes, Nurses' Health Study, Nurses' Health Study II, and Health Professionals' Follow-up Study, The National Institute of Neurological Disorders and Stroke Genetics Network, Netherlands Twin Register, Australian Twin-Family Studies on Nicotine and Alcohol Genetics, SardiNIA project), UK Biobank, and Women's Health Initiative | Age at which an individual started smoking cigarette regularly measured by several ways including "At what age did you begin smoking regularly?" and "How long have you smoked?" combined with "What is your current age?". | $n = 262,990$ |
| Smoking cessation<br>(Liu et al., 2019) | Self-report | Meta-analysis performed from Add Health, Avon Longitudinal Study of Parents and Children, Atherosclerosis Risk in Communities, Biobank Japan Project, The Barrett's and Esophageal Adenocarcinoma. Genetic Susceptibility Study, Brisbane Longitudinal Twin Study, Center on Antisocial Drug Dependence, Collaborative Genetic Study of Nicotine Dependence, Genetics of Chronic Obstructive Pulmonary Disease, deCODE Genetics/AMGEN, Inc., Estonian Genome Center, Electronic Medical Records and Genomics, Finnish Twin Cohort, Framingham Heart Study, Genetic Epidemiology Research in Adult Health and Aging, Genes for Good, The Nord-Trøndelag Health Study, Health and Retirement Study, Minnesota Center for Twin and Family Research, Multi-Ethnic Study of Atherosclerosis, Metabolic Syndrome in Men, Netherlands Study on Cognition, Environment and Genes, Nurses' Health Study, Nurses' Health Study II, and Health Professionals' Follow-up Study, The National Institute of Neurological Disorders and Stroke Genetics Network, Netherlands Twin Register, Australian Twin-Family Studies on Nicotine and Alcohol Genetics, SardiNIA project), UK Biobank, and Women's Health Initiative | Current smokers coded as "2" and former smokers coded as "1", and never smokers are coded as missing. | $n = 312,821$ |
| Cigarettes per day<br>(CPD)<br>(Liu et al., 2019) | Self-report | Meta-analysis performed from Add Health, Avon Longitudinal Study of Parents and Children, Atherosclerosis Risk in Communities, Biobank Japan Project, The Barrett's and Esophageal Adenocarcinoma. Genetic Susceptibility Study, Brisbane Longitudinal Twin Study, Center on Antisocial Drug Dependence, Collaborative Genetic Study of Nicotine Dependence, Genetics of Chronic Obstructive Pulmonary Disease, deCODE Genetics/AMGEN, Inc., Estonian Genome Center, Electronic Medical Records and Genomics, Finnish Twin Cohort, Framingham Heart Study, Genetic Epidemiology Research in Adult Health and Aging, Genes for Good, The Nord-Trøndelag Health Study, Health and Retirement Study, Minnesota Center for Twin and Family Research, Multi-Ethnic Study of Atherosclerosis, Metabolic Syndrome in Men, Netherlands Study on Cognition, Environment and Genes, Nurses' Health Study, Nurses' Health Study II, and Health Professionals' Follow-up Study, The National Institute of Neurological Disorders and Stroke Genetics Network, Netherlands Twin Register, Australian Twin-Family Studies on Nicotine and Alcohol Genetics, SardiNIA project), UK Biobank, and Women's Health Initiative | Cigarettes per day responses were binned as<br>a. 1=1-5<br>b. 2 = 6-15<br>c. 3 = 16-25<br>d. 4 = 26-35<br>e. 5=36+ | $n = 263,954$ |
| Lifetime cannabis use (Pasman et al., 2018) | Self-report | UK Biobank and International Cannabis Consortium | Defined as any use of cannabis during lifetime | $n = 162,082$ |
| Cannabis Use Disorder (Johnson et al., 2020) | Clinician ratings or semi-structured interviews | Psychiatric Genomics Consortium Substance Use Disorders working group, The Integrative Psychiatric Research, and deCODE | Met criteria for a lifetime diagnosis of DSM-IV (or DSM-III-R) cannabis abuse or dependence, DSM-5 cannabis use disorder and ICD-10 codes of F12.1 (cannabis abuse) or F12.2 (cannabis dependence). | $n = 384,032$ |
| Opioid Use Disorder (Zhou et al., 2020) | Clinician ratings | Million Veteran Program, Yale-Penn, and Study of Addiction: Genetics and Environment | International Classification of Diseases, Ninth Revision-diagnosed OUD or International Statistical Classification of Diseases and Related Health Problems, Tenth Revision-diagnosed OUD (Million Veteran Program), and DSM-IV-defined opioid dependence (Yale-Penn and Study of Addiction: Genetics and Environment). | $n = 114,759$ |

#### Substance Use Measures

Our analysis used summary statistics from several of the largest GWAS of substance use behaviors. Alcohol behaviors included alcoholic drinks per week (DPW) ^58^ and problematic alcohol use (PAU) ^43^. Tobacco behaviors included lifetime tobacco use, age of first becoming a regular smoker, smoking cessation, and cigarettes per day (CPD) ^58^. Cannabis behavior included lifetime cannabis use ^59^ and Cannabis Use Disorder (CUD) ^60^. Lastly, we included Opioid Use Disorder (OUD) ^47^. All trait discovery cohorts were of European ancestry. Table 1 details each substance use trait in terms of type of measure, measure construct, coding, and discovery sample.

### Analyses

#### Linkage Disequilibrium Score Regression

We used Linkage Disequilibrium Score Regression (LDSC) ^61^ to estimate genetic correlations between traits. GWAS summary statistic SNPs were removed if they had MAF > 0.01 and INFO > 0.70. SNPs, duplicated rs numbers, were multi-allelic variants, were strand ambiguous, had deletions/insertions or had low Ns. Alleles were merged with the Hap Map 3 ^62^ reference panel (major histone complex removed). Beta weights and linkage disequilibrium (LD) were pre-generated from 1000 Genomes European GWAS data included in the LDSC software download. LDSC regresses Chi-square statistics from the summary stats of GWAS on LD scores of the trait of interest ^61^. LD scores for each SNP are calculated via the sum of the variance explained by LD of that SNP with other SNPs ^63^. Genetic correlations were estimated using overlapping SNPs from filtered summary statistic files provided from GWAS summary statistics. LDSC accounts for possible sample overlap and additional sources of confounding (e.g., population stratification). We estimated a pair-wise genetic correlation matrix that included all sleep/activity and substance use measures (figure 7). Due to the large number of genetic correlations estimated, we utilized Bonferroni correction ^64^ to adjust for potential false positives.

#### K-means Clustering of correlations

K-means clustering is an unsupervised machine learning clustering technique that uses a centroid or distance-based algorithm to assign correlations to a cluster of a predefined number ^65, 66^. After assigning the K number of clusters, the algorithm shuffles the data to clusters and assigns them to initial random centroids. It then determines the sum of squares (or distance) between each data point and the initial centroids and does a series of reassignments to the centroids until the algorithm is finished with the appropriate clusters, attempting to make data in the clusters similar while making each individual cluster separate from the others. By comparing the distance (sum of squares) within clusters across different number of clusters (i.e. 1 vs. 2 and 2 vs. 3) we can estimate a silhouette coefficient that shows the best number of clusters to account for patterns in the data, i.e. the cluster solution that allows for the smallest sum of squares across clusters.

An advantage of K-means over other clustering algorithms is that clustering is done at the variable level with the correlation matrix (unlike mixture modeling or DBSCAN) in a hypothesis free format (unlike GenomicSEM). The latter is necessary for this analysis as large GWAS studies do not share individual data. For the current analyses, the genetic correlation matrix was read into the K-Means algorithm. The silhouette coefficient determined how many centers were needed to keep each substance use and sleep measure closest together, compared to other potential cluster solutions. K-means clustering was conducted in R using the R packages “cluster”^67^ and “factoextra”^68^.

#### Latent Causal Variable Analysis

To examine evidence for genetic causality between sleep and substance use/misuse phenotypes, we used latent causal variable analysis (LCV) ^69^ on genetic correlations that survived Bonferroni correction. LCV allows genetic correlations between two traits to be mediated by a latent variable with a causal effect on each trait. This model was designed to account for genetic pleiotropy by partitioning the genetic correlation into pleiotropy vs partial causality. This is done using the 4^th^ order moments from LD Score Regression^69^. Using this model, causality is implied when trait one is strongly correlated with the causal latent variable compared to the second trait, suggesting that part of the genetic component of trait one is causal for the second trait in the relationship. If trait one is perfectly genetically correlated with the latent variable, it can be considered fully genetically causal. The extent to which the latent causal variable causes trait 1 versus trait 2 is expressed as a ratio, referred to as the genetic causality proportion (gcp). The gcp is an estimate of the degree to which each trait is correlated with the latent genetic variable with a score that can range from 0 (reflecting no genetic causality) to 1/-1 (signifying full genetic causality). For instance, a gcp of 0.70 would suggest 70% of SNP effect sizes are consistent with trait 1 causing trait 2. It is worth noting that LCV estimates the direction of causality in terms of the order of variables, if the gcp is positive, trait 1 is genetically causal for trait 2. If the gcp is negative, trait 2 is genetically causal for trait 1. For the current analyses, the sleep/activity measure is trait 1 and the substance use measure is trait 2.

While LCV is similar to other methods of determining genetic causality in that it utilizes SNPs to derive instrumental variables (like traditional Mendelian randomization), LCV has advantages over other methods of estimating genetic causality. First, sample overlap is accounted for by the LDSC intercept. Second, the model produces a genetic causality proportion, that is the proportion of genetic effects that are consistent with a model of genetic causality, allowing us to account for partial overlap. Third, the gcp is robust to pleiotropy, which is accounted for using 4^th^ order moments from LDSC. Fourht, the pleiotropic effects across the entire genome are accounted for, rather than only at a few SNPs^69^. Indeed, simulation designs shows that the method outperforms Mendelian randomization^69^ when the genetic correlation between traits is non-zero. Considering that many of our samples overlap and that we want to account for pleiotropy, LCV is ideal over MR in this analysis.

## Results

### Genetic Correlations Across Sleep and Substance Use

Figure 1 displays the genetic correlation matrix between all substance use behaviors and sleep/activity measures. We found 31 significant pair-wise correlations across domains that survived Bonferroni correction (Table 2), with several trends suggesting strong genetic relationships between substance use behaviors and sleep/activity measures. In all cases, greater vulnerability for substance use was associated with more sleep problems. Measures of both subjective self-reported sleep duration (long, full, and short) and objective accelerometer derived sleep duration had genetic correlations with numerous substance use traits including lifetime tobacco use, age of first becoming a regular smoker, CPD, smoking cessation, PAU, lifetime cannabis use, CUD, and OUD (absolute values of rGs between 0.11 - 0.32). Increased diurnal inactivity was genetically correlated with lifetime tobacco use, age of initiation of smoking, CPD, and DPW (Absolute values of rGs between 0.12 – 0.18), Insomnia was also genetically correlated with numerous substance use traits including lifetime tobacco use, age of initiation of regular smoking, CPD, smoking cessation, DPW, PAU, CUD, and OUD (absolute values of rGs between 0.11-0.36).

**Figure 1.**
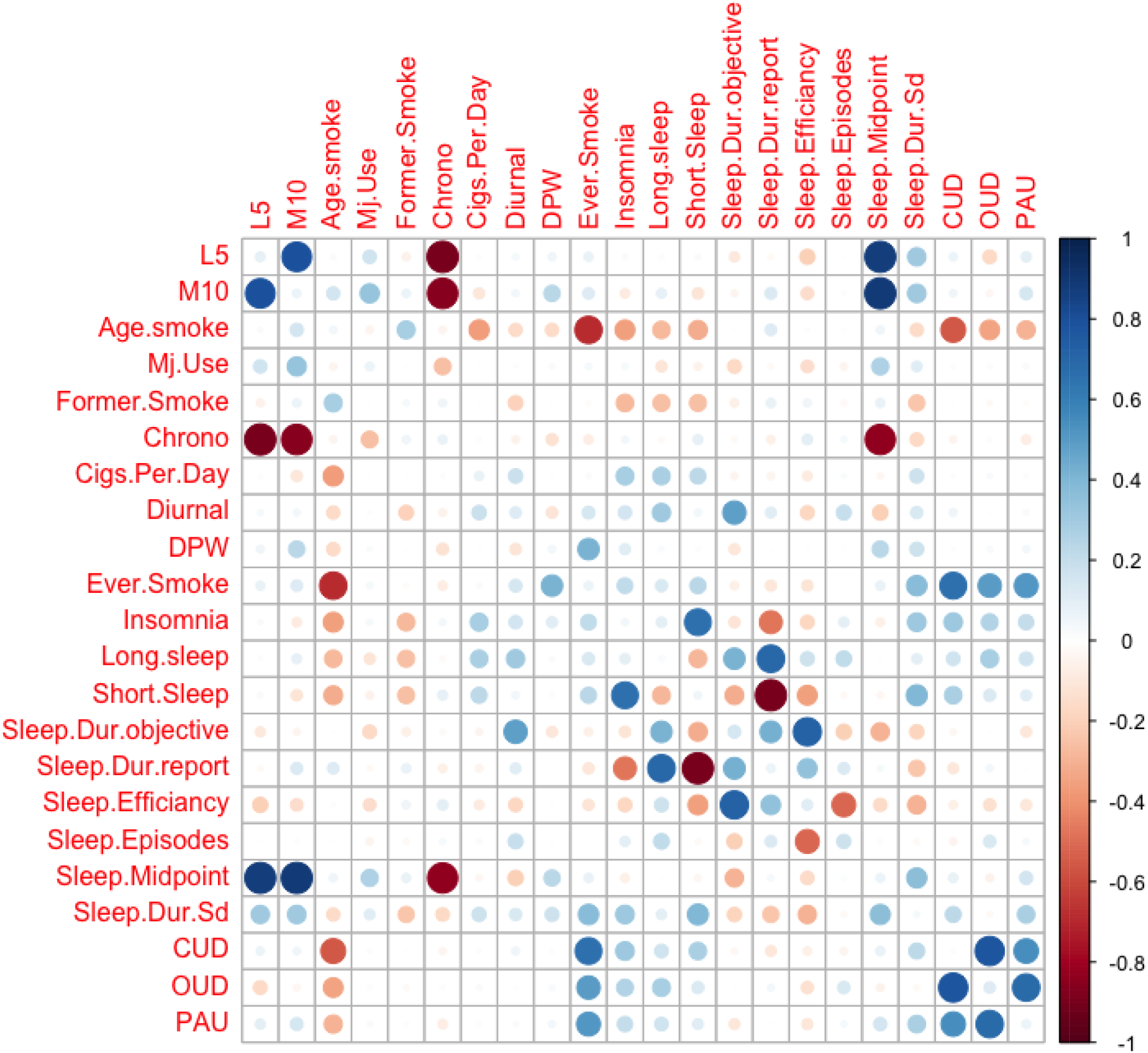
The genetic (rG) correlation matrix between all substance use behaviors and sleep/activity measures. Positive rGs shown in blue and negative in red. Correlations are plotted on X and Y axis based of centroid clustering. Age. Smoke = Age of first becoming a regular smoker, CPD = Cigarettes per week, Former.Smoke = current smoker versus formal smoker, DPW= Drinks per week, OUD= Opiate Use Disorder, CUD = Cannabis Use Disorder, PAU = Problematic Alcohol Use, Ever.Smoke = lifetime tobacco use, Long sleep = Subjective self-report short sleep duration, Short Sleep = Subjective self-report short sleep duration, Sleep.Dur.Sd = Accelerometer Sleep Duration (Standard Deviation), Sleep.Dur.report = subjective self-report sleep duration.

**Table 2.**
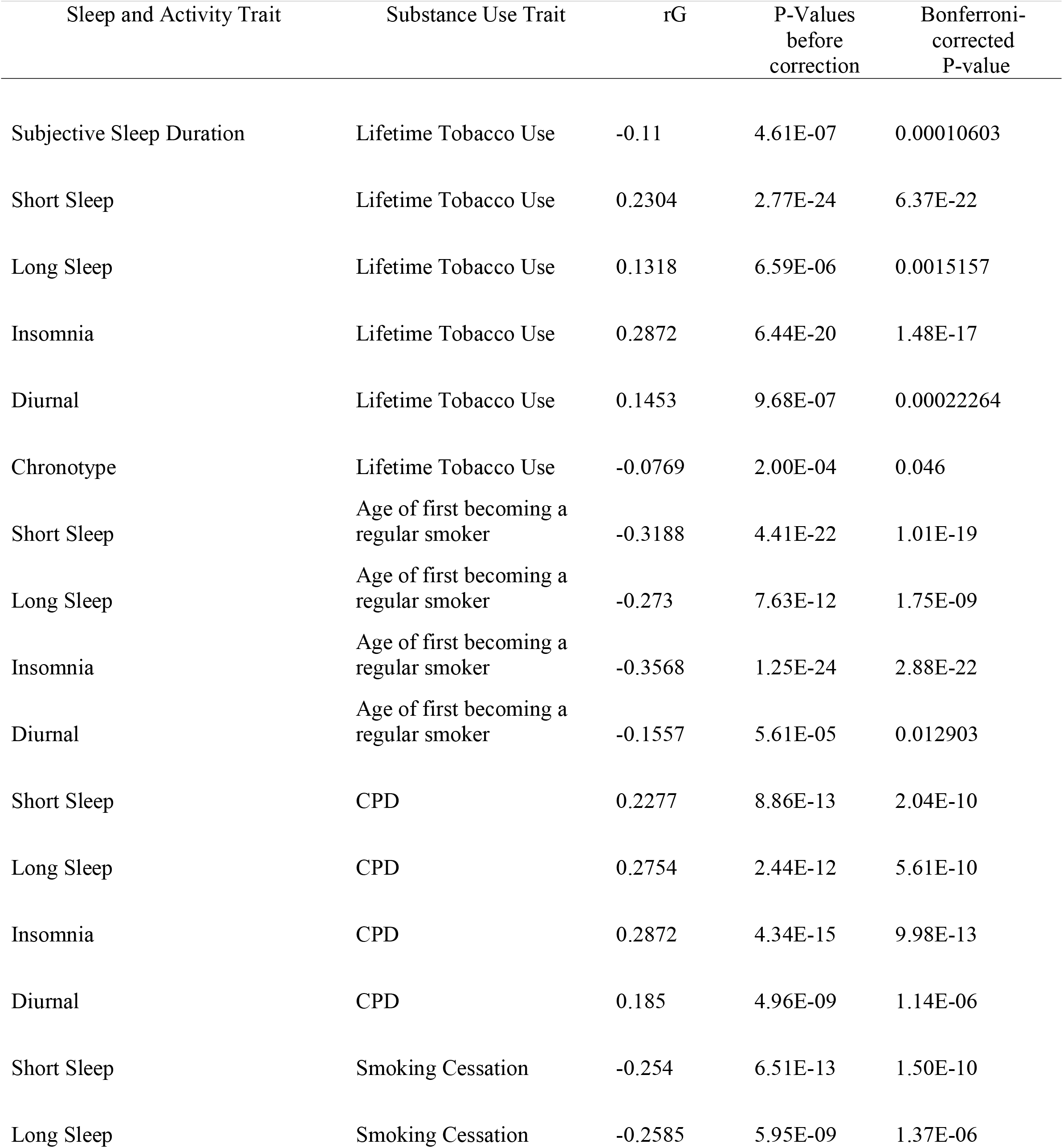
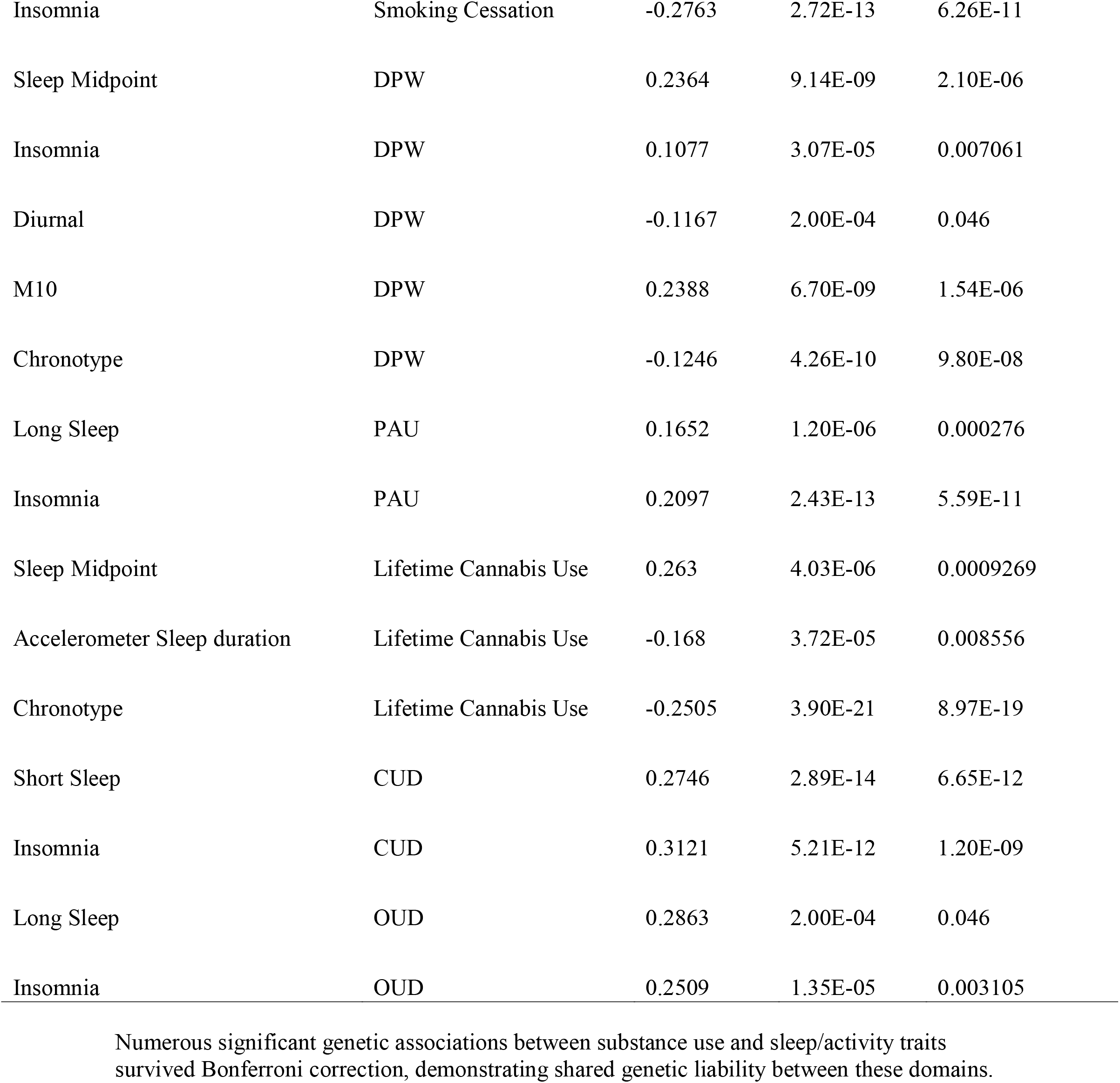
Significant pair-wise correlations that survived Bonferroni multiple corrections between substance use and sleep/activity traits.

Only a few of the genetic correlations involving the activity based circadian-related endophenotype measures survived Bonferroni corrections including 1) sleep midpoint with lifetime cannabis use (rG = 0.26), and DPW (rG = 0.24) and 2) M10 with DPW (rG = 0.24). Focusing on measures of chronotype, in addition to the circadian-related sleep midpoint relationships mentioned above, significant genetic correlations were found between self-reported chronotype and lifetime cannabis use, lifetime tobacco use, and DPW (Absolute values of rGs between 0.08 – 0.25).

### K-means clustering within and between sleep and substance use domains

A silhouette coefficient determined that 2 clusters were optimal to explain the overlap between sleep/circadian-related activity and substance use dimensions (Figure 2). Figure 3 displays our optimal cluster solution. We refer to the first cluster as the “tobacco use severity” cluster, which grouped the substance use behaviors of age of initiation of regular smoking and smoking cessation with elements of sleep health such as self-report long sleep duration, self-report sleep duration, accelerometer derived sleep duration, sleep efficiency, and self-report chronotype. The traits most central to this cluster included sleep efficiency (sleep duration divided by the time between the start and end of the first and last nocturnal inactivity period) and self-reported sleep duration. The second cluster reflected “substance use and use disorders” and contained common substance use behaviors (lifetime tobacco use and lifetime cannabis use), consumption behaviors (CPD and DPW) as well as problematic substance use behaviors (CUD, OUD, and PAU) with measures of sleep difficulties (insomnia, self-report short sleep duration, increased number of sleep episodes, increased standard deviation of accelerometer derived sleep duration, and diurnal inactivity) and circadian-related activity measures (L5, M10, and sleep midpoint). The traits most central to this cluster included DPW, PAU, and the standard deviation of accelerometer derived sleep duration.

**Figure 2.**
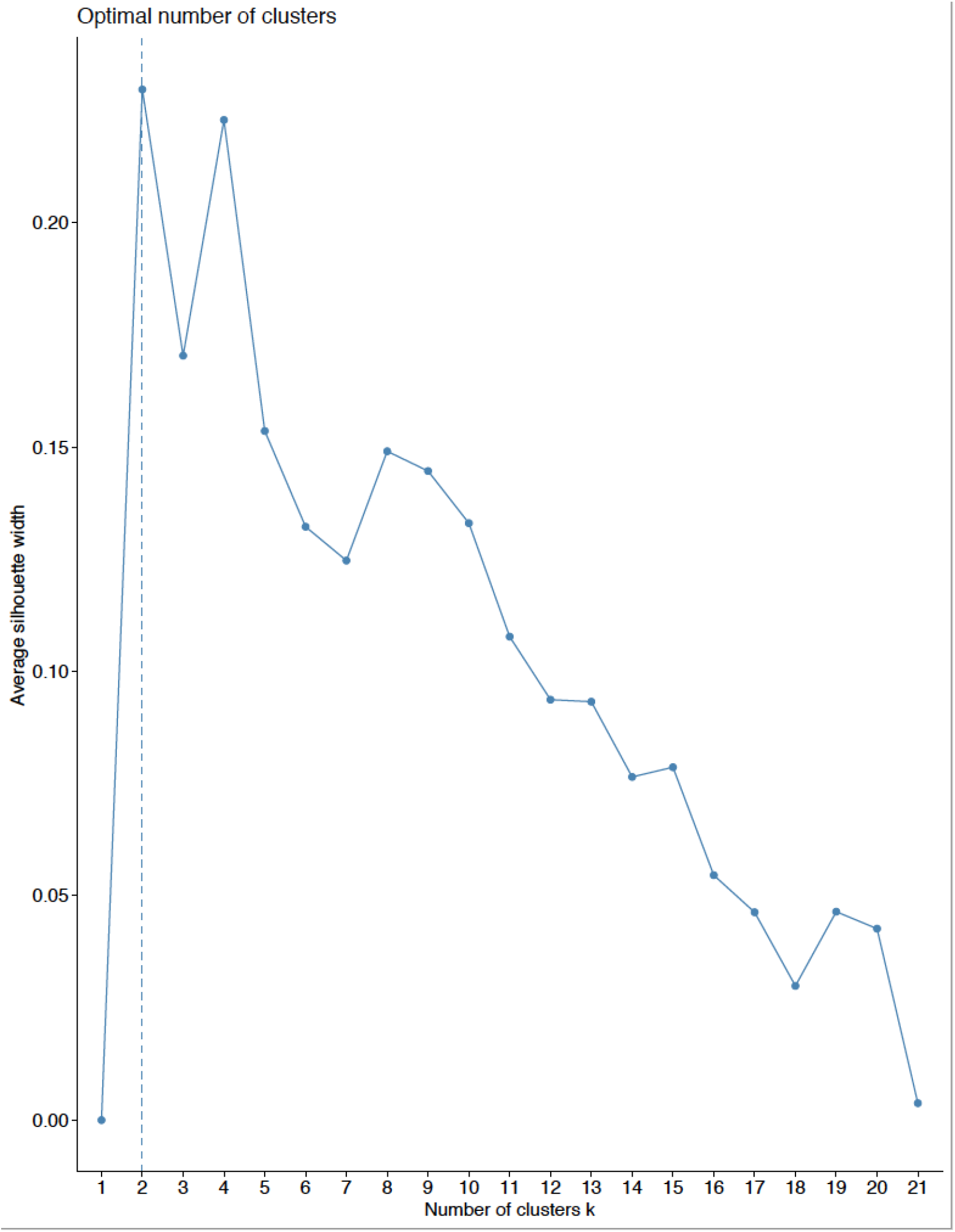
Silhoutte Coefficents from K-Means Clusting Algorithm A silhouette coefficient determined that a 2-cluster solution was optimal to characterize the relationships between sleep/activity and substance use dimensions.

**Figure 3.**
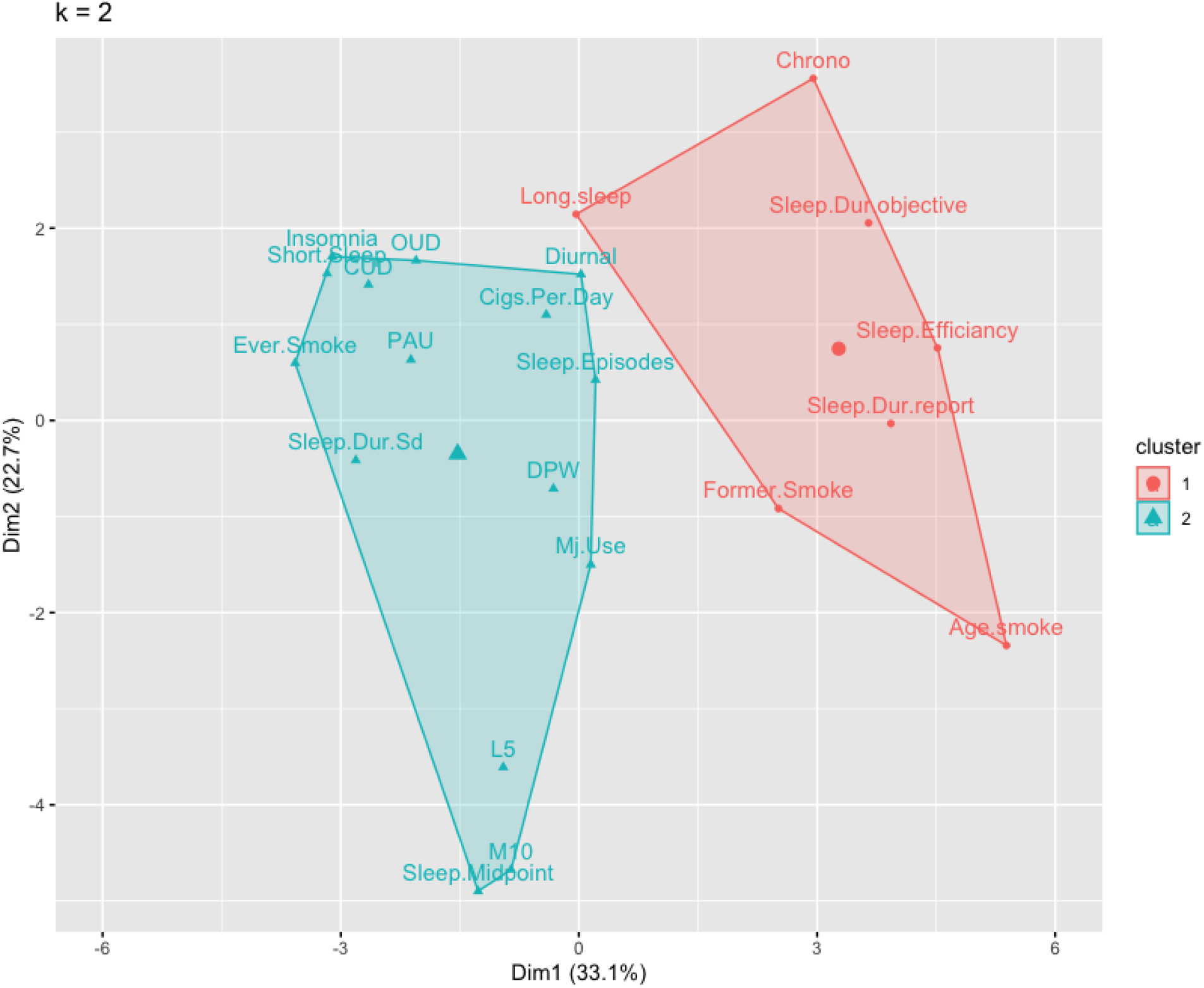
Optimal cluster solution demonstrated two distinct clusters, both of which were comprised of common substance use behaviors and sleep traits. Clusters are plotted by a two dimensional representation of the derived components. X and Y axis represent the first two components and their proportion of variance they explain in each trait. Clusters are plotted around their centroid. Smoke = Age of first becoming a regular smoker, CPD = Cigarettes per week, Former.Smoke = current smoker versus formal smoker, DPW= Drinks per week, OUD= Opiate Use Disorder, CUD = Cannabis Use Disorder, PAU = Problematic Alcohol Use, Ever.Smoke = lifetime tobacco use, Long sleep = Subjective self-report short sleep duration, Short Sleep = Subjective self-report short sleep duration, Sleep.Dur.Sd = Accelerometer Sleep Duration (Standard Deviation), Sleep.Dur.report = subjective self-report sleep duration.

### Latent Causal Variable Analysis

We used latent causal variable analysis between each of the 31 substance use and sleep/activity measure genetic correlations that survived Bonferroni correction. Of the 31 LCV models, one model survived further Bonferroni correction among the LCV models (.05/31=.0016), with evidence of OUD being genetically causal for self-report long sleep duration (gcp = 0.50, corrected *p* = 0.01). Several models reached nominally significant p-values before Bonferroni correction, including CUD with subjective short sleep duration (gcp = −0.23, nominal *p* = 0.04), CUD with insomnia (gcp = −0.29, nominal *p* = 0.05), and OUD with insomnia (gcp = −0.35, nominal *p* = 0.05). A majority of the models lacked significance before correction, indicating a lack of support for causality between substance use/misuse and sleep measures (Table 3). Thus, there was scarce evidence of genetic causality once pleiotropy, polygenicity, and sample overlap are appropriately accounted for.

**Table 3.** Results from latent causal variable analysis to test causality between associated sleep/activity and substance use dimensions.

| Sleep and Activity Trait | Substance Use Trait | GCP (standard Error) | P-values before correction | Bonferroni-corrected P-values |
| --- | --- | --- | --- | --- |
| Self-report Short Sleep Duration | Lifetime Tobacco Use | -0.14(0.10) | 0.10 | 1 |
| Insomnia | Lifetime Tobacco Use | -0.08(0.10) | 0.32 | 1 |
| Self-report Full Sleep Duration | Lifetime Tobacco Use | -0.20(0.28) | 0.16 | 1 |
| Diurnal inactivity | Lifetime Tobacco Use | 0.12(0.29) | 0.39 | 1 |
| Self-report Long Sleep Duration | Lifetime Tobacco Use | -0.37(0.37) | 0.39 | 1 |
| Chronotype | Lifetime Tobacco Use | -0.26(0.41) | 0.25 | 1 |
| Insomnia | Age of first becoming a regular smoker | -0.10(0.11) | 0.39 | 1 |
| Self-report Short Sleep Duration | Age of first becoming a regular smoker | -0.03(0.08) | 0.79 | 1 |
| Self-report Long Sleep Duration | Age of first becoming a regular smoker | -0.33(0.19) | 0.16 | 1 |
| Diurnal inactivity | Age of first becoming a regular smoker | 0.13(0.26) | 0.50 | 1 |
| Insomnia | CPD | 0.31(0.41) | 0.63 | 1 |
| Self-report Short Sleep Duration | CPD | -0.21(0.20) | 0.32 | 1 |
| Self-report Long Sleep Duration | CPD | -0.06(0.27) | 0.63 | 1 |
| Diurnal inactivity | CPD | -0.41(0.35) | 0.32 | 1 |
| Insomnia | Smoking Cessation | -0.43(0.27) | 0.13 | 1 |
| Self-report Short Sleep Duration | Smoking Cessation | 0.01(0.12) | 0.79 | 1 |
| Self-report Long Sleep Duration | Smoking Cessation | -0.10(0.19) | 0.63 | 1 |
| Chronotype | DPW | -0.05(0.21) | 0.63 | 1 |
| M10 | DPW | -0.32(0.21) | 0.25 | 1 |
| Sleep Midpoint | DPW | -0.45(0.32) | 0.32 | 1 |
| Insomnia | DPW | -0.21(0.47) | 0.50 | 1 |
| Diurnal inactivity | DPW | 0.04(0.53) | 0.63 | 1 |
| Insomnia | PAU | -0.08(0.12) | 0.50 | 1 |
| Self-report Long Sleep Duration | PAU | 0.03(0.28) | 1 | 1 |
| Chronotype | Lifetime Cannabis Use | 0.15(0.16) | 0.39 | 1 |
| Sleep Midpoint | Lifetime Cannabis Use | 0.21(0.38) | 0.63 | 1 |
| Accelerometer derived sleep duration | Lifetime Cannabis Use | -0.05(0.46) | 0.16 | 1 |
| Self-report Short Sleep Duration | CUD | -0.23(0.11) | 0.04 | 1 |
| Insomnia | CUD | -0.29(0.14) | 0.05 | 1 |
| Insomnia | ODU | -0.35(0.17) | 0.05 | 1 |
| **Self-report Long Sleep Duration** | **ODU** | **-0.50(0.25)** | **0.00036** | **0.01116** |

Significant models that survived Bonferroni correction in bold

## Discussion

Using summary data from the largest publicly available GWAS of both sleep/circadian-related activity and substance use measures to date, we found 31 significant genetic correlations between traits in these domains that survived Bonferroni correction. Clustering analysis uncovered two principal genetic clusters: 1) the tobacco use severity cluster and 2) the substance use and use disorders cluster. Latent causal variable analyses confirmed that the associations between sleep/activity measures and substance use behaviors were driven primarily by common or shared genetic influences, with one exception, a model which implied that OUD may plausibly exert a genetically causal influence on self-reported long sleep duration. The results herein suggest a strong pleiotropic genetic relationship between the domains of substance use and sleep/circadian-related activity measures.

We found significant genetic correlations that align with prior findings, including genetic relationships between subjective sleep measures and substance use behaviors involving cannabis ^48^, tobacco^44, 45^, alcohol^46^, and opioids^47^. We also found novel relationships between subjective measures of sleep and substance use, such as OUD with subjective long sleep duration. Of primary interest are the genetic correlations involving substance use and objective measures of sleep and circadian-related activity, most of which have not been reported previously. For example, genetic associations between lifetime cannabis use and objectively measured later sleep mid-point and decreased accelerometer derived sleep duration are the first reports of a genetic relationship between cannabis use and objective measures in this domain of research. Later sleep midpoint is an objective proxy for an evening chronotype ^26, 54, 55^, and these results corroborate prior subjective findings regarding a genetic relationship between lifetime cannabis use and self-report evening chronotype ^48^. These findings suggest a shared genetic relationship of lifetime cannabis use with circadian-related measures, implying that people who have more evening or nighttime activity are more likely to have used cannabis due to a shared genetic vulnerability.

While a prior result of objective sleep duration variation and lifetime tobacco use were not replicated ^56^, this is the first report of significant genetic correlations between numerous tobacco behaviors and subjective sleep measures. These results imply shared genetic underpinnings between a range of increased tobacco behaviors (lifetime use, earlier age of first becoming a regular smoker, increased cigarettes per day, and smoking cessation) with both problematic objective sleep duration (short and long sleep duration) and increased diurnal inactivity during the day. These relationships suggest the genetics responsible for increased tobacco use are also responsible for improper sleep duration and the potential sleep compensation/rest that can accompany. Genetic correlations between drinks consumed (DPW) and sleep midpoint, diurnal inactivity, and M10 are the first report of a genetic relationship between alcohol use and objective measures of sleep and circadian-related activity. Interestingly, increased DPW being genetically correlated with less diurnal inactivity, later midpoint (proxy for evening chronotype), and increased M10 (more active later in the day) imply that genetic predisposition to increased drinking might also be associated with increased activity throughout the day and being more active later in the day or evening. Thus, these differential associations may be of interest in developing sleep-assisted interventions, such that adjusting chronotype could be a novel intervention for issues related to alcohol use.

These genetic correlations formed two distinguishable and well-delineated clusters. The first cluster suggests that genes associated with aspects of tobacco use severity (a younger age of regular smoking initiation and smoking cessation) share genetic overlap with elements of sleep health such as sleep duration (self-report long sleep duration, shorter self-report sleep duration, and shorter accelerometer derived sleep duration), sleep efficiency, and self-report chronotype. Sleep health is an established research term that provides a frame of reference for the field ^70^ and includes sleep elements such as sleep duration, efficiency, and chronotype. These sleep health elements clustering together imply a genetic relationship between indicators of heavy tobacco use and sleep health. Tobacco use is highly comorbid with insufficient sleep ^71^ and it is speculated that the relationship could be centered on potential nightly withdrawals, the strong stimulant effects of nicotine, and an increased prevalence of sleep disordered breathing in comparison to nonsmokers ^72^. Exploration of sleep deficits in tobacco withdrawal may benefit from understanding shared genetic vulnerabilities to poor sleep health in particular.

The second cluster found that substance use behaviors, such as consumption (lifetime tobacco use, lifetime cannabis use, DPW, and CPD) and problematic use (CUD, OUD, and PAU) clustered strongly with measures of problematic sleep (insomnia, self-report short sleep duration, increased number of sleep episodes, increased standard deviation of accelerometer derived sleep duration, and diurnal inactivity) as well as measures of circadian-related activity (L5, M10, and sleep midpoint). Insomnia deficits are thought to be part of the “dark side” of substance use ^73^ that reflects negative reinforcement-related drug intake, and seems to manifest specifically with diagnosis (and likely heavy use) based on this cluster. Interestingly, recent drug targets for these negative affect/withdrawal-related symptomology of addiction have been shown to improve insomnia symptoms as well. For instance, Acamprosate that is used to treat alcohol use disorder targets withdrawal symptoms and thus improves sleep symptoms during treatment_74_.

Though some previous work argues the association between insomnia and substance use is causal (via Mendelian randomization) ^49^, based on results herein, these findings are likely confounded by pleiotropy. Our approach accounted for pleiotropy and did not confirm the previous findings, although we did find one novel association. Specifically, our LCV analyses provided novel evidence that liability to OUD may be genetically causal for self-report long sleep duration, even after accounting for pleiotropy between these traits. Over 80% of individuals with OUD report poor sleep quality and sleep problems which likely impedes opioid maintenance and other pharmacological treatments ^75, 76^. Opioid therapy for pain and related conditions has been well-documented to disrupt sleep and due to its respiratory depressant effects, exacerbate risk for sleep-disordered breathing ^77^. While this is the first finding of a significant relationship between OUD and long sleep duration, long sleep duration has been found to be phenotypically correlated with depression, antidepressant use, benzodiazepine use, and heavy drinking ^78^, as well as genetically correlated with depression ^52^. Thus, the causal genetic relationship demonstrated could reflect more of an underpinning of the genetics associated with a general maladaptive behavioral profile in comparison to specific opiate use genetics. While three other models were nominally significant before correcting for multiple tests, the lack of significance in these models ultimately implies pleotropic inferences as the more supportive explanation of the associations. Thus, it is worth noting that pleiotropy may underlie sleep problems in substance use, and that populations entering substance use treatment are also likely to be at greater vulnerability for sleep deficits.

## Limitations

There are several limitations of this study that would influence LDSC and LCV analyses and the generalizability of our findings. First, GWAS results are comprised of mostly common variants; therefore, no rare variation would be included in our analyses, but rare variants may be an additional source of genetic overlap or provide stronger instruments for causality. Second, the GWAS summary statistics used in our analyses assume that the genomic liability to a trait is a good instrument for manifesting the phenotype (for example, genomic liability for insomnia is a good predicter of having insomnia), but this may not necessarily be true for all traits. Third, LCV and LDSC are best when used in one ancestral population, and all GWAS traits were of European ancestry. There is a need to include more racially diverse and multi-racial samples in genetics research. Finally, the objectively measured sleep/activity phenotypes/endophenotypes (the accelerometer derived traits) had much smaller samples than the self-report sleep measures; this difference in statistical power may mean that genetic correlations with the accelerometer traits were less likely to be statistically significant that those for self-report sleep measures, and therefore less likely to be carried forward in the causal analyses.

## Conclusions

While substances such as cannabis and alcohol are often used as sleep aids, individuals with substance use disorders also struggle with sleep difficulties, and poor sleep complicates pathways to sustained remission from substance use disorders. Our study documents the role of shared genetic influences on substance use disorders and both sleep and circadian-related measures indicating specific domains of overlap that may be used to discover mechanisms of intervention. For opioid use disorder in particular, mechanisms of association likely extend beyond pleiotropy into potential causality. Together, these results imply a strong shared genetic relationship between the domains of common substance use behaviors and sleep traits.

## Data Availability

All data in this study is publicly available. See Table 1 for information on each sample.

## Acknowledgements and Funding

ASH receives support from DA007261-17. EAW receives support from T32 MH015442 and DA017637. CLM receives support from MH016880. ECJ receives support from K01DA051759.

